# Concurrent of stunting and overweight/obesity among children: evidence from Ethiopia

**DOI:** 10.1101/2020.02.13.20022723

**Authors:** Alinoor Mohamed Farah, Tahir Yousuf Nour, Bilal Shikur Endris, Seifu Hagos Gebreyesus

**Affiliations:** Department of Public Health, College of Medicine and Health Sciences, Jigjiga University, Ethiopia; Department of Reproductive Health and Health Service Management, School of Public Health, College of Health Sciences, Addis Ababa University, Ethiopia

**Author notes:** Corresponding Author Correspondence and reprint requests should be addressed to: Alinoor Mohamed Farah, College of Medicine and Health Sciences, Jigjiga University, Jigjiga, Ethiopia, Lecturer at Jigjiga University.

**Keywords:** Young children, Concurrent of Stunting and Overweight/Obesity, Stunting, Overweight, DHS, Ethiopia

## Abstract

**Background:** Nutrition transition in many low- and middle-income countries (LMICs) has led to childhood nutritional outcomes to shift from a predominance of undernutrition to a dual burden of under- and overnutrition. Yet, Infant and young child feeding programs in Ethiopia mainly focus on undernutrition. It is therefore crucial to assess the prevalence and determinants to better inform infant young child feeding programs.

**Methods:** We analyzed anthropometric, sociodemographic and dietary data of children aged 6-23 months from 2016 Ethiopian Demographic and Health Survey (EDHS). A total of 2,674 children were included in the current study. Concurrent of Stunting and Overweight/Obesity (CSO) prevalence was estimated by distal, intermediate and proximal factors. To identify factors associated with CSO, we conducted hierarchical logistic regression analyses.

**Results:** The overall prevalence of CSO was 2.45%. The odds of CSO was significantly higher in children of low household wealth category, compared to their counter parts in the richest category (AOR=2.07, 95%CI=1.06–4.03, P=0.033). In boys, the likelihood of CSO was 1.60 times that of girls (95%CI =1.03–2.49, P=0.038). Above 12months of age was significantly associated with a higher odd of CSO, compared to below 12months of age (AOR=1.76, 95% CI=1.07– 2.88, P=0.026).

Small birth size was significantly associated with a higher odd of CSO, compared to large birth size (AOR=2.53, 95%CI=1.45–4.41, P=0.001). Children who did not take vitamin A supplement within the previous six months had a higher odd of CSO, compared to those who took (AOR=1.91, 95%CI =1.19–3.07, P =0.007).

**Conclusion:** Our study provided evidence on the co-existence of stunting and overweight/obesity among infants and young children in Ethiopia. CSO was associated with various factors originating from community and child levels. Therefore, identifying children at risk of growth flattering and excess weight gain provides IYCF programs in Ethiopia and beyond with an opportunity of earlier interventions.

## Introduction

Child malnutrition which includes both undernutrition and overweight are global challenges which is associated with an increased risk of mortality and morbidity, unhealthy development, and loss of productivity[1]. Although improvement of child undernutrition in Ethiopia has been achieved, stunting remains an important problem in Ethiopia, with thirty eight percent of children <5 years of age affected. However, economic growth and urbanization in countries like Ethiopia have given rise to a nutrition transition, where there is as shift from traditional diets to “western diets” (energy-dense diets)[2]. This has led to an increase in overweight and obesity. On the other hand, undernutrition decreased but the decline has not been happening as quickly as the rise in overweight and obesity, leading to a double burden of overnutrition and undernutrition[3].

Dual burden of malnutrition could occur at country, household, or individual levels. At household level at least one member is undernourished and at least one member is overweight whereas at individual level the dual burden of malnutrition often manifests as stunting or micronutrient deficiencies co-occurring with overweight or obesity[3-5]. At individual level, history of stunting coupled with consumption of high dense energy foods and micronutrient deficiencies owing to shared underlying determinants or physiologic links may also result in clustering of nutrition problems such as co-occurrence of stunting and overweight particularly among young children[3].

Children who are stunted and gain abnormal weight later can be at greater risk of unhealthy development than other children. In other words they are at high risk of non-communicable disease since they impose a high metabolic load on a depleted capacity for homoeostasis[6]. Further, concurrent under and over nutrition it a public health challenge in a sense there is a need to strike a delicate balance between reducing undernutrition and preventing over nutrition [5]. There are substantial studies on dual burden of malnutrition and mainly focused on prevalence and trends of double burden at different levels[2-5, 7-28] but few studies investigated factors associated with the dual burden at individual level and in particular among infant and young children [13, 18, 21, 22, 25].

To the best of our knowledge there were no previous studies in Ethiopia on cooccurrence of stunting and overweight among infants and young children that explored factors associated with it. The previous studies focused on inclusive measure of child undernutrition and failed to capture the severity of malnutrition for some children who suffer from more than one type of malnutrition. Therefore, aim of this study is to assess the prevalence of co-occurrence of stunting and overweight/obese among infants and young children in Ethiopia and factors associated with it.

## Study subjects and Methods

### Study subjects

We analyzed anthropometric data of children aged 6-23 months from 2016 EDHS which is nationally representative survey. A total of 2925 children aged 6–23 months were found in the dataset. A total of 2674 children were extracted and included in the final dataset after exclusion of incomplete data.

A multistage stratified two-stage cluster sampling procedure was used to select samples. A detailed information of sample design and procedure is presented elsewhere [29]. We included children aged 6-23 months with complete height, weight, dietary and non-dietary data.

### Study design and data source

We used data set from national representative cross-sectional household survey that was conducted from September to December 2015. At individual level, we estimated the prevalence of double burden of malnutrition defined as co-existence of stunting and overweight/obese within the same child. Stunting was defined as length-for-age Z-score (LAZ) below -2SD and overweight/overweight was defined as BMI-for-age Z-score (BAZ) above 2SD from the respective WHO 2006 growth standards reference median. Weight were measured using SECA scales while length was measured using Shorr measuring boards and children were measured while lying down [29].

The data used for this study was extracted from the fourth EDHS. The survey was implemented by Central Statistical Agency (CSA) at the request of Ethiopian Federal Ministry of Health (FMoH) and funded by United States Agency for International Development (USAID).

### Data analysis

Children recode data file in the form of STATA was used for analysis. Statistical analysis was performed using the STATA software package, version 14.1 (Stata Corp., College Station, TX, USA). Survey command (svy) was used to adjust for the complex sample design.

We estimated weighted prevalence of CSO by distal, intermediate and proximal factors. Overall differences across the categories were statistically tested using design-based Pearson chi-squared test. Multiple hierarchical logistic regression was used to examine the effect of distal, intermediate and proximal factors. First bivariate analyses were done for all potential predictors of CSO. Then, hierarchical regression models were run using variables which demonstrated P<0.20 during the bivariate analyses. The three-level hierarchical regression models were run following the recommendation of a previous study that suggested to take into account complex hierarchical relationships of different determinants at different level [30]. The first, second, and third models included distal, intermediate, and proximal factors, respectively. As mentioned before, variables with P<0.20 value during the bivariate analyses were included in the multiple hierarchical logistic regression analyses. In other words, model-1, 2 and 3 included the distal, intermediate and proximal factors which demonstrated P<0.20 during the bivariate analyses. To put it differently, we used model-1 to assess the overall effect of distal factors and excluded the intermediate and proximal factors. We used model 2 to assess the effect of intermediate factors in the presence of distal factors which were considered confounding factors. Proximal factors were entered in model 3 in the presence of distal and intermediate factors which were also considered confounding variables in model 3. Variable significant at p-value of 0.05 during the hierarchical regression analyses was considered to be determinant factor at each model in which the variable was first entered regardless of its performance in the subsequent model(s). The approach was meant to avoid the possibility of underestimating the effects of distal factors[30].

## Result

### Background characteristics

The background characteristics of children are presented in Table 1. Fifty three percent of children were females and the rest were male. Sixty six percent of children were older than twelve□months and the rest were younger than twelve months. Overall, 31.4 and 11.9% of children were stunted and overweight, respectively. The overall prevalence of CSO was 2.45%. CSO prevalence among urban and rural children was 1.62 and 2.56%, respectively. Among boys and girls, the prevalence was 2.96 and 2.0%, respectively. The age-specific estimates were 1.86 and 2.77% in those aged under 12 months and above 12 months, respectively. The prevalence of CSO by other child characteristics is shown in Table 1.

**Table 1:** Bivariate analysis of the relation of distal, intermediate, and proximal factors with CSO

|  |  | Weighted<br>frequency<br>(%) | CSO prevalence<br>(95% CI) | *P*-<br>value* |
| --- | --- | --- | --- | --- |
| *Basic factors*<br>*(distal)* | Urban | 11.6 | 1.62 [0.41, 6.14] | 0.52 |
| Residence<br>place | Rural | 88.4 | 2.56[1.78, 3.67] |  |
| Wealth | Low | 44.3 | 3.28[2.08, 5.14] | 0.27 |
|  | Middle | 22.9 | 1.94 [0.77, 4.81] |  |
|  | High | 32.8 | 1.67[0.79, 3.52] |  |
| Caregiver<br>education | No | 62.2 | 3.31[2.23, 4.89] | 0.02 |
|  | Primary | 31.1 | 0.68[0.26, 1.78] |  |
|  | Secondary+ | 6.7 | 2.64[0.63, 10.34] |  |
| *Underlying*<br>*factors*<br>*(intermediate)* |  |  |  |  |
| Water source<br>type | Not<br>improved | 42.1 | 2.26[1.3, 3.88] | 0.72 |
|  | Improved | 57.9 | 2.59[1.61, 4.15] |  |
| Toilet facility<br>source | Not<br>improved | 90.6 | 2.34[1.64, 3.33] | 0.56 |
|  | Improved | 9.4 | 3.48[0.95, 11.94] |  |
| *Immediate*<br>*factors*<br>*(proximal)* |  |  |  |  |
| Child sex | Boy | 46.9 | 2.96[1.82, 4.78] | 0.30 |
|  | Girl | 53.1 | 2.00[1.16, 3.41] |  |
| Child age | <12 months | 34.4 | 1.83[0.81, 4.08] | 0.37 |
|  | >12 months | 65.6 | 2.77[1.86, 4.11] |  |
| Birth size | Large | 31.4 | 1.86[0.81, 4.22] | 0.07 |
|  | Average | 40.8 | 1.71[0.87, 3.34] |  |
|  | Small | 27.8 | 4.25[2.26, 6.99] |  |
| Infection<br>history | No | 62.6 | 2.12[1.27,3.50] | 0.35 |
|  | Yes | 37.4 | 3.00[1.80,2.95] |  |
| Breastfeeding<br>duration | <12 months | 36.0 | 1.92[0.90, 4.06] | 0.41 |
|  | >12 months | 64.0 | 2.75[1.85, 4.07] |  |
| Early<br>breastfeeding<br>initiation | No | 15.3 | 4.37[2.22, 8.43] | 0.07 |
|  | Yes | 84.7 | 2.10[1.36, 3.23] |  |
| Deworming | No | 91.2 | 2.18[1.47, 3.24] | 0.09 |
|  | Yes | 8.80 | 5.16[2.06, 12.38] |  |
| Vitamin A<br>supplement | No | 60.3 | 2.99[1.98, 4.50] | 0.14 |
|  | Yes | 39.7 | 1.62[0.79, 3.27] |  |
| Iron<br>supplement | No | 92.3 | 2.57[1.79, 3.68] | 0.06 |
|  | Yes | 7.7 | 0.96[0.35, 2.63] |  |
| Grains | No | 39.1 | 1.93[1.03, 3.59] | 0.35 |
|  | Yes | 60.9 | 2.78[1.81, 4.26] |  |
| Fruits and vegetables | No | 70.2 | 2.71[1.82, 4.02] | 0.39 |
|  | Yes | 29.8 | 1.82[0.81, 4.07] |  |
| Meat | No | 91.7 | 2.66[1.86, 3.78] | <0.001 |
|  | Yes | 8.3 | 0.13[0.00025,0.67] |  |
| Diary | No | 82.9 | 2.20[1.44, 3.35] | 0.25 |
|  | Yes | 17.1 | 3.64[1.75, 7.41] |  |
| Pulses | No | 79.8 | 2.55[1.73, 3.76] | 0.61 |
|  | Yes | 20.2 | 2.03[0.90, 4.51] |  |
| Eggs | No | 83.8 | 2.50[1.70, 3.66] | 0.80 |
|  | Yes | 16.2 | 2.18[0.80,5.82] |  |
| MDD | No | 87.4 | 2.49[1.71, 3.62] | 0.80 |
|  | Yes | 12.6 | 2.14[0.67, 6.49] |  |
| MMF | No | 57.9 | 1.60[0.92, 2.78] | 0.03 |
|  | Yes | 42.1 | 3.61[2.24, 5.78] |  |

CSO: Concurrent of stunting and overweight/obesity CI: confidence interval MDD: Minimum Dietary Diversity MFF: Minimum meal frequency \* based on chi-square test of association

### Determinants of CSO

The multiple hierarchical logistic regression model presented in Table 2 indicated that house hold wealth, child’s age and sex, birth size and vitamin A supplementation were significantly associated CSO.

**Table 2:**
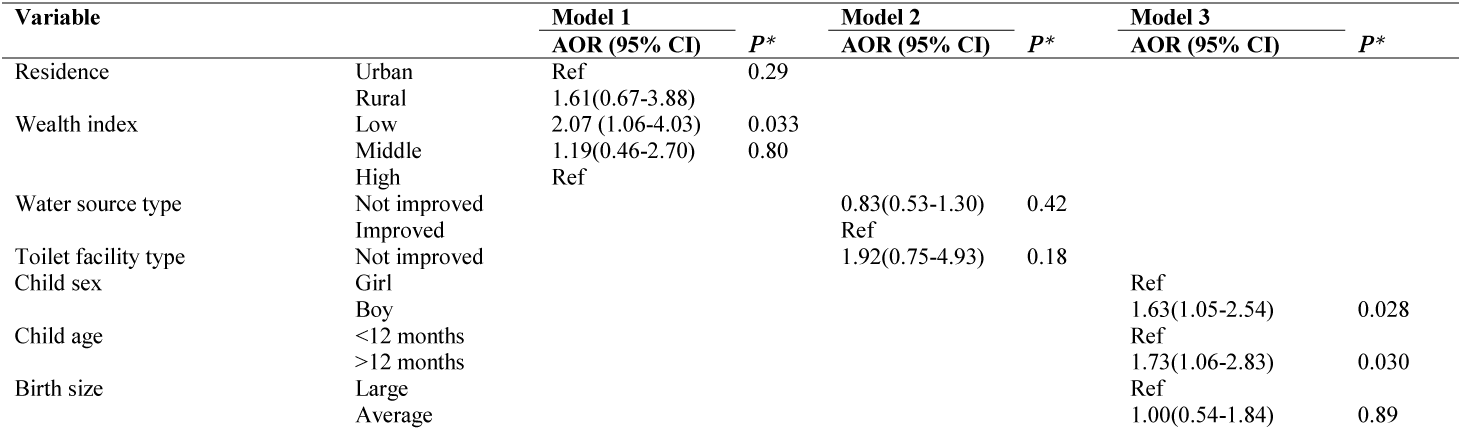
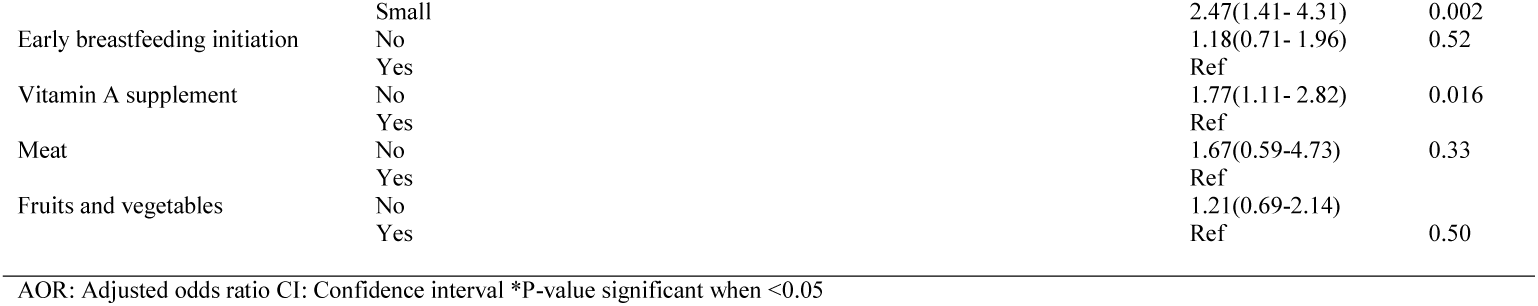
Hierarchical multiple logistic regression analysis of the relation of basic, underlying, and proximal factors with CSO

The odds of CSO was significantly higher in children of low household wealth category, compared to their counter parts in the richest category (AOR=2.07, 95%CI=1.06–4.03, P=0.033). In boys, the likelihood of CSO was 1.60 times that of girls (95%CI=1.03–2.49, P =0.038). Above 12months of age was significantly associated with a higher odd of CSO, compared to below 12months of age (AOR=1.76, 95% CI=1.07– 2.88, P=0.026).

## Discussion

Using a national representative sample of infants and young children, the current study aimed to determine the prevalence of CSO and its associated factors among infants and young children in Ethiopia. Our study provided evidence that there was co-occurrence of stunting and overweight among infants and young children in Ethiopia. We found CSO associated with factors at different levels. The basic factor associated with higher odds of CSO was being poor. The immediate factors found associated with higher odds of CSO were male sex, age above 12 months, small birth size and no vitamin A supplement use for the last six months. Our results showed high level of stunting among infants and young children, with 31.4% of children being stunted. Overweight was also prevalent, though not as high as stunting. We found that the 2.45% of children concurrently suffered stunting and overweight/obese. There are studies that documented prevalence of co-occurrence of stunting and overweight among under five children in African countries. The prevalence determined by the current study was higher than studies compared to previous study conducted in Ghana that assessed dual burden of malnutrition among children younger than five years [22] but lower than a study conducted in South Africa and Libya that reported prevalence of 18% and 7% respectively among under five children [15, 27].

Similarly, studies from Asia and Latin America countries determined the prevalence of concurrent of stunting and overweigh. The prevalence of this report was within the range of report from Ecuador that reported a prevalence of 2.8%[10], higher than study from Mexico that reported a prevalence of 1%[16] and lower than a prevalence from China that reported 5.06%[21]. A recent study that examined the dual burden of malnutrition among children aged 6-59 months in the Middle-East and North Africa (MENA) and Latin American and Caribbean (LAC) regions also showed a prevalence that ranged from 0.4 to 10.7% in MENA regions and 0.3 to 1.9% in LAC regions [31]. The prevalence of current study is higher than prevalence of almost all LAC regions and within the range in most countries in MENA regions.

We also found several factors associated with CSO. Children of low household wealth were more likely to be concurrently stunted and overweight/obese. Children of low household wealth were more likely to be concurrently stunted and overweight/obese. This could be due to the fact that infant and young child caring practices like hygiene, proper feeding and health services utilization are often poorly practiced among poor households compared to their richer counterparts[32-34]. This finding also agrees with the existing literature on concurrent of stunting and overweight [22, 25]. Children who reside in the rural areas had a higher risk of being concurrently stunting and overweight though insignificant. This finding was also consistent with the existing literature which shows children residing in rural and less urbanized areas are at risk of being concurrently stunted and overweight[21].

We also found higher odds of CSO in boys and those children above 12 months of age. This could be due to fact that most stunting occurs more during the period 12 to 23months of age[32, 35]. In general, children under-two years of age bear a higher burden of both overweight and stunting, particularly in LMICs countries[21]. Further, our finding of higher risks of CSO in boys than in girls was not in agreement with previous reports which demonstrated higher odds of stunting and overweight in girls [22]. Small birth size was also significantly associated with CSO. This finding was also in agreement with the existing evidence which suggests low birthweight linked to poor health and nutritional outcomes [1, 35, 36]. Fetal growth restriction is an important contributor to stunting in children and evidence showed that low birthweight was associated with 2.5–3.5-fold higher odds of wasting, stunting and underweight[36]. Further, other evidences have suggested low birth weight babies who exhibit catchup growth may be at risk of abnormal weight gain in childhood[37-41]. In other words, intra uterine growth retardation might induce a catch-up fat mass for the fetus before even born [42] and the rapid weight gain appears to promote adiposity after about age 2 years particularly in LMICs [43]. Thus, it could be easily acknowledged that low birth weight child would be at a higher risk of being concurrently affected by stunting and overweight/obesity. Vitamin A intake was also associated with a significantly lower CSO prevalence. This would be most likely due to the role of vitamin A in promoting linear growth and reduce the risk of stunting[44-47].

The first 1,000 days is a critical time for physical and intellectual growth and set a foundation for long term heath and development [1]. As a result of greater awareness of significance of stunting as one of major public health problems, stunting reduction has gained increased international attention [48]. Similarly, overweight/obesity in this age group deserves attention because at early stage of life catch up growth have been identified as one of the risk factors that lead to progression of abnormal weight gain. [49-55]. Therefore, it is important to identify children that are at risk of developing CSO as early as possible to limit progression of both growth flattering and abnormal weight gain. Further, evidence have shown that if only under nutrition is targeted with an aim of targeting growth may unintentionally contribute to abnormal weight gain[56].

The current study has the following limitations; first, recall bias while reporting the birth, infection, dietary history of children is still an issue of concern [57]. In other words, collecting data like birth size, history of infection and dietary related data are solely based on memory of mothers or the caretaker which might have led to recall bias. Further, due to the cross-sectional nature of this study a cause and effect relationship could not be inferred.

Given the above-mentioned limitations, the current work has some strengths. First, the EDHS data is a national representative data and conclusions about Ethiopia can be drawn. Second the data is reliable and of high quality since the standardized procedures are employed by such kind of survey. Third, appropriate statistical method was used to explore the relationships between the outcome variable and its determinants.

## Conclusion

In conclusion, our study provided evidence on the co-existence of stunting and overweight/obesity among infants and young children in Ethiopia. CSO was associated with various factors originating from community and child levels. Therefore, identifying children at risk of growth flattering and excess weight gain provides IYCF programs in Ethiopia and beyond with an opportunity of earlier interventions.

## Data Availability

All data are fully available without restriction

## Abbreviations

AOR: Adjusted Odds Ratio
BAZ: Body Mass Index for Age Z-score
BMI: Body Mass Index
CI: Confidence Interval
CSO: Concurrent of Stunting and Overweight/Obesity
EA: Enumeration Area
EDHS: Ethiopia Demographic and Health Survey
IYCF: Infant and Young Child Feeding
LAZ: Length for Age Z-score
IRB: Institutional Review Board
LAC: Latin American and Caribbean
LMIC: Lower, Middle Income Countries
MAD: Minimum Acceptable Diet
MENA: Middle-East and North Africa
MDD: Minimum Dietary Diversity
MMF: Minimum Meal Frequency
NRERC: National Research Ethics Review Committee
SGA: Small for Gestational Age UNICEF United Nations Children’s Fund
USAID: United States of Agency for International Development
WHO: World Health Organization

## Declarations

## Acknowledgment

The authors will like to thank ICF international to grant permission to use the EDHS data.

## Availability of data and materials

The dataset for this study is available from the corresponding author on reasonable request.

## Ethical approval

Ethical clearance for the survey was provided by Institutional Review Board (IRB) of the College of Medicine and Health Sciences at Jigjiga University. Online application to analyze the secondary data was requested from DHS Program, USAID and we have been authorized to download data from the Demographic and Health Surveys (DHS) online archive.

## Consent for publication

Not applicable

## Authorship

AM conceived the study, prepared the proposal, analyzed the data, interpreted the findings and wrote the manuscript. TY, BS and SH were involved in developing the study proposal, data analysis and reviewing the manuscript.

